# Disease-driven domain generalization for neuroimaging-based assessment of Alzheimer’s disease

**DOI:** 10.1101/2023.09.22.23295984

**Authors:** Diala Lteif, Sandeep Sreerama, Sarah A. Bargal, Bryan A. Plummer, Rhoda Au, Vijaya B. Kolachalama

## Abstract

Development of deep learning models to assess the degree of cognitive impairment on magnetic resonance imaging (MRI) scans has high translational significance. Performance of such models is often affected by potential variabilities stemming from independent protocols for data generation, imaging equipment, radiology artifacts, and demographic distributional shifts. Domain generalization (DG) frameworks have the potential to overcome these issues by learning signal from one or more source domains that can be transferable to unseen target domains. We developed an approach that leverages model interpretability as a means to improve generalizability of classification models across multiple cohorts. Using MRI scans and clinical diagnosis obtained from four independent cohorts (Alzheimer’s Disease Neuroimaging Initiative (ADNI, *n* = 1, 821), the Framingham Heart Study (FHS, *n* = 304), the Australian Imaging Biomarkers and Lifestyle Study of Ageing (AIBL, *n* = 661), and the National Alzheimer’s Coordinating Center (NACC, *n* = 4, 647)), we trained a deep neural network that used model-identified regions of disease relevance to inform model training. We trained a classifier to distinguish persons with normal cognition (NC) from those with mild cognitive impairment (MCI) and Alzheimer’s disease (AD) by aligning class-wise attention with a unified visual saliency prior computed offline per class over all training data. Our proposed method competes with state-of-the-art methods with improved correlation with postmortem histology, thus grounding our findings with gold standard evidence and paving a way towards validating DG frameworks.

## 1 INTRODUCTION

Alzheimer’s disease (AD) is a progressive syndrome leading to loss of brain function that affects memory, thinking, language, judgment and behavior. The approach to dementia diagnosis involves careful consideration of the patient’s demographics and symptoms, family, social and medical history, neurologic examination, cognitive, behavioral, and functional assessments along with neuroimaging [1, 2]. Magnetic resonance imaging (MRI) is typically recommended to evaluate the structural changes in the patient’s brain that correspond to volume loss and atrophy patterns suggestive of AD and rule out other patterns indicative of non-AD syndromes. Computational methods based on advanced machine learning techniques are increasingly considered to automatically process the MRI scans and classify persons with AD dementia from those with normal cognition (NC) and mild cognitive impairment (MCI) [3, 4, 5, 6]. Some of recently reported frameworks have relied on training models using data collected from a single cohort followed by evaluation on independent test cohorts [6]. Such model development strategies can establish a proof-of-principle, but may lack generalizability because data collected from multiple cohorts may contain variabilities stemming from independent scanning protocols, diversity of the study population and other sources. Moreover, while recent efforts on public data sharing have facilitated data access more easily, there is a greater need to develop models that result in generalizable, consistent findings.

Recently, domain generalization (DG) approaches are being considered to train robust deep learning models that account for cohort-specific variabilities and work well across multiple datasets [7, 8, 9, 10, 11, 12, 13, 14, 15]. Most methods attempt to mitigate the distributional variance between domain-specific feature representations. We submit that additional aspects such as orienting the models to focus on disease-related information while performing model training can be a targeted approach to meet the objective of creating generalizable architectures for disease classiffication.

### 1.1 Related work

DG frameworks are typically designed to learn a robust signal and a set of patterns possibly from single or multiple source domains with the aim of transferring them to unseen target domains. The expectation is that such frameworks lead to minimal performance degradation on the unseen target environment. In the setting of single-source DG, the model trained on this source learns robust representations that can generalize to out-of-distribution data. Single-source DG methods can also be applied to a multi-source setting, as training is done over pooled data across the different source domains [8]. Also, multiple source domains can be used for training domain-invariant feature representations that generalize well to unseen target data.

Most DG methods were originally designed to benchmark natural imaging datasets, with a limited number of frameworks focused on medical imaging data [9, 10]. A group of methods have been proposed to tackle DG via data manipulation, which could either be data augmentation or generation [16, 17, 18, 19, 20]. One of those methods is Mixup[16], a data-agnostic routine that constructs virtual training examples as convex combinations of pairs of examples and their labels sampled at random from the training distribution. Mixup aims to regularize the neural network to favor linear behavior in-between training examples[16]. Another group of methods used representation learning to address domain shift, mainly by learning domain-invariant representations and feature disentanglement [11, 12, 21, 22, 13, 15, 23]. Donini and co-workers proposed a multi-source algorithm that uses empirical risk minimization (ERM), which became the standard approach to the DG problem. ERM aims to minimize the training risk across all source domains[11]. Recently, Kreuger and colleagues introduced risk extrapolation (REx) for out-of-distribution (OOD) generalization and proposed a penalty on the variance of training risks (V-REx). They showed that reducing differences in risks with V-REx can reduce a model’s sensitivity to a wide range of extreme distributional shifts[12]. Li et al. (2018), on the other hand, proposed using the maximum mean discrepancy (MMD) measure with autoencoders to align distributions across different domains via adversarial training [13]. Another work introduced representation self-challenging (RSC) to force the model to discard dominant features activated on the training data and activate remaining features that correlate with ground-truth labels [15]. Further, there exists a line of work that used meta-learning for DG. One of the proposed meta-learning methods was MLDG, meta-learning for domain generalization, which simulates domain shift during training by synthesizing virtual testing domains within each mini-batch [14].

Our method is orthogonal to previous work on learning domain-invariant feature representations, with the contribution of using interpretability to extract disease-relevant information for feature alignment. Related prior work used model explanations as means of disentangling domain specific information from otherwise relevant features[24]. Our method, on the other hand, leverages feature contributions to final correct predictions as prior knowledge of model-identified disease biology to guide model attention during training. We focus on the single-source DG setting, as it is the more feasible scenario in clinical settings, where the model has access to only one source domain for training. The model’s generalizability is then evaluated on external cohorts known as the target domains.

### 1.2 Contributions

Our work falls under the umbrella of interpretability, where we use feature contributions to adjust final predictions and emphasize disease-relevant features. Through attention-based supervision, the model learns to focus on disease-correlated regions using precomputed class-wise saliency map priors with voxel contributions. The main contributions of this paper are summarized as follows:

- We developed an interpretability-based computational framework to train deep neural networks that focus on model-identified disease regions of interest as a means to generalize across multiple cohorts.
- Using MRI scans and clinical data obtained from multiple cohorts, we developed a classifier that distinguishes between persons with NC, MCI and AD.
- We demonstrated that our method competes with state-of-the-art DG methods in the real-world single-source setting.
- Finally, we showed that our interpretable findings correlate strongly with postmortem histology, identifying disease presence in brain regions that are known to classically associate with disease.

## 2 METHODS

### 2.1 Study population

We obtained brain MRI scans and corresponding clinical and demographic data on participants from four different cohorts: the Alzheimer’s Disease Neuroimaging Initiative (ADNI) (*n* = 1, 821) [25], National Alzheimer’s Coordinating Center (NACC) (*n* = 4, 647)[26], the Australian Imaging Biomarkers and Lifestyle (AIBL) study of ageing (*n* = 661) [27], and the Framingham Heart Study (FHS) [28, 29] (*n* = 304). There were 3, 697 cases with normal cognition (NC), 2, 323 cases with mild cognitive impairment (MCI), and 1, 413 cases with Alzheimer’s disease (AD) across all cohorts (Table 1). The selection criterion included T1-weighted 1.5 and 3 Tesla MRI scans of individuals aged 55 years and older taken within ±6 months from the date of clinically confirmed diagnosis [6]. We considered the MRI scan taken at the closest date to the diagnosis when multiple MRIs were available per person. Any cases including AD with mixed dementia, non-AD dementias, history of severe traumatic brain injury, depression, stroke, and brain tumors, as well as incident major systemic illnesses were excluded. We considered the single-source setting for DG, where training, internal validation and testing of the models were performed on one source cohort, whereas external validation and testing were performed on the target cohorts.

**T A B L E 1.**
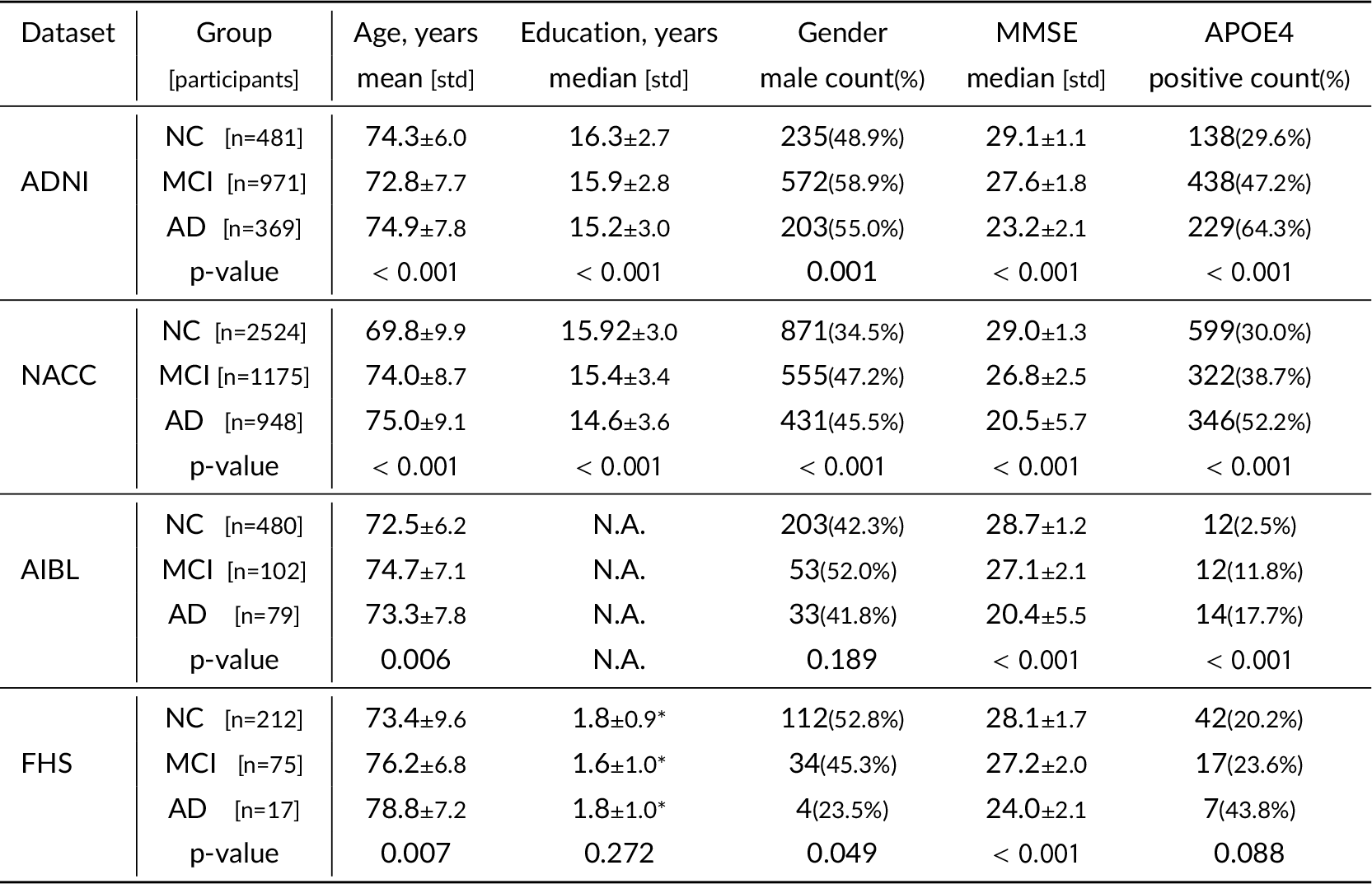
Study population. MRI scans and corresponding clinical and demographic data were collected across four different cohorts: the Alzheimer’s Disease Neuroimaging Initiative (ADNI), the National Alzheimer’s Coordinating Center (NACC), the Australian Imaging, Biomarker & Lifestyle Flagship Study of Ageing (AIBL), and the Framingham Heart Study (FHS). The models were trained and tested to differentiate persons who have either normal cognition (NC), mild cognitive impairment (MCI) or Alzheimer’s disease dementia (AD). Education information on the AIBL dataset was not available. ^*^FHS education code: 0 = high school did not graduate, 1 = high school graduate, 2 = some college graduate, 3 = college graduate.

### 2.2 MRI processing pipeline

We uniformly applied a series of image processing steps to all the MRI scans. First, we reconfigured the scans’ orientation to follow that of the MNI space. We then used the FSL brain extraction tool to generate a mask that identified brain voxels comprising gray and white matter, cerebrospinal fluid, and subcortical structures, including the brain stem and cerebellum. In this step, any pixels corresponding to white matter, cerebrospinal fluid, brain stem and cerebellum were removed. Following skull stripping, we applied a preliminary linear registration of the MRIs to the Montreal Neurological Institute (MNI-152) coordinate system. Skull stripping and linear registration were once again applied to remove any brain tissue outside of the full registered brain and enhance alignment with the template. Then, intensity artifacts were removed via bias field correction to increase data homogeneity. Finally, a nonlinear warp of the Hammersmith Adult brain atlas was applied to the post-processed MRIs to obtain the parcellated brain regions. The above preprocessing steps were done following Qiu *et al*. (2022) [30].

### 2.3 Computational framework

We developed our computational framework for the task of classification of 3D volumetric brain scans into NC, MCI, and AD. The building blocks of our framework are a feature extractor, a class-wise attention module, and a classifier network (Fig. 1). The training pipeline consists of two stages: the first is training a baseline model for the offline computation of class-wise priors, and the second is training a new independent model with the supervision of these priors.

**F I G U R E 1.**
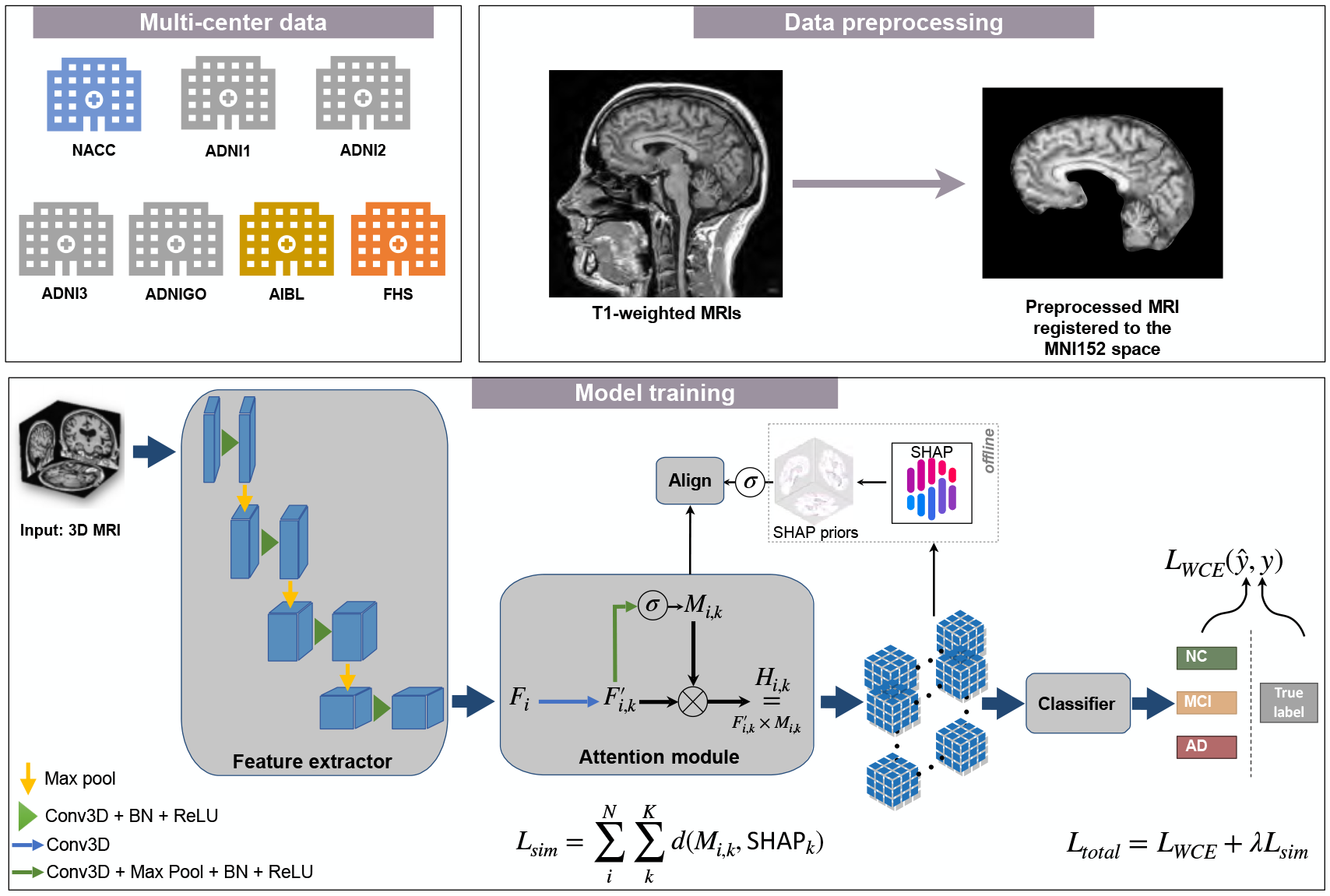
Schematic of the disease-informed domain generalization framework. MRI scans from various cohorts were processed via an image processing pipeline that was previously described [6]. Our approach takes 3D MRIs as input from the source domain and learns their feature representations in the latent space, and uses an attention module to learn class-specific saliency maps. These maps are then used to predict a class label (NC, MCI, or AD). We used SHAP offline to generate the averaged saliency maps, which we refer to as disease-informed prior knowledge, of NC, MCI, and AD classes over all samples of the source domain used for model training.

#### Feature extractor

We chose the UNet3D [31] architecture and started from a pretrained `Models Genesis` checkpoint on chest CT scans [32, 33]. `Models Genesis` are generic pre-trained 3D models for 3D medical image analysis. They were trained in a self-supervised robust manner, and outperformed models trained from scratch [33]. To adapt the network to our classification task, we discarded the decoder module and kept the encoder of the UNet3D network as the feature extractor for our framework.

#### Classifier module

We used a global average pooling (GAP) layer[34] followed by a softmax function as the classifier for the three-way classification of NC, MCI, and AD. Our choice of a GAP layer as opposed to a fully connected layer as the classifier encourages spatial awareness. While the latter takes in as input a feature map pooled over the channel dimension and then flattened into a one-dimensional vector, the former receives a stack of 3D feature maps with the channel dimension *K* being equivalent to the number of classes, and pools over the spatial dimensions thereby preserving spatial information per channel.

#### Attention supervision

We added an attention module between the feature extractor and the classifier to learn class-wise attention over the source domain. It takes as input the feature maps *F*_*k*_ output by the feature extractor, and passes it through a 3D convolutional layer to get 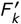. The learnt attention maps are denoted by *M*_*k*_ ∈ℝ^*K* ×*D* ×*H* ×*W*^, where *K* is the number of classes, and *D, H*, and *W* are the depth, height, and width of the attention map, respectively. The final output of the attention module is then the element-wise multiplication of 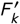 and *M*_*k*_ . The class-wise attention maps were later used in the second stage of training for alignment with visual saliency priors computed per class over the training data.

#### Training

In the first phase of training, we computed visual saliency maps over correct predictions by a baseline model trained with weighted cross-entropy over the training data. To achieve this task, we used SHapley Additive exPlanations (SHAP) to compute the feature contributions per class [35]. For the purpose of smoothing out sample noise and variance, we used an averaged saliency map over samples of the same class as a representation of class-wise saliency. Fig. 2 shows visualizations of the pre-computed SHAP prior specific to the AD class. For the purpose of visualization, Shapley values were scaled to the range of [™1, 1], which we chose to correctly represent negative and positive voxel contributions as in the original range. Once the SHAP priors were generated, we ran our explainability-based strategy to regularize the model through a combined weighted cross entropy (1) and similarity loss (2). We applied augmentation techniques to the training data using the Medical Open Network for AI (MONAI) framework [36], which included random contrast adjustment, random bias field, random spatial cropping, upsampling, and intensity scaling. We found that intensity scaling to the range [0, 1] worked best for data normalization of structural MRI scans.

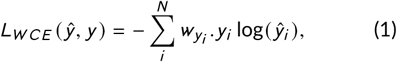

such that *N* spans the minibatch dimension, and 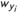 refers to the weight assigned to all samples belonging to the ground-truth class *y*_*i*_ . Class weights are computed by taking the inverse of the total count of samples belonging to each class, so that underrepresented classes have a higher weight.

**F I G U R E 2.**
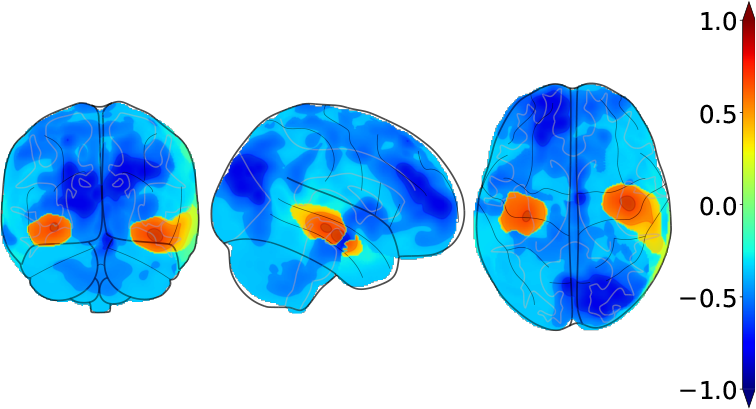
Orthogonal projections of the pre-computed AD-specific SHAP prior used in our computational framework. The above projections correspond to the averaged saliency maps with respect to correct predictions of AD over all samples of the source domain. We projected the resulting map to 2D space onto the coronal, sagittal, and axial axes, respectively.

After having the SHAP maps generated offline per class, we used a similarity loss defined in (2) to minimize the distance between each sample’s extracted feature maps and the retrieved SHAP prior with respect to the same class as the ground truth label of that sample.

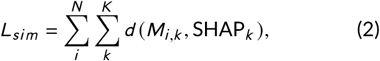

with *d* being the distance metric of choice, which, in our case, is the L2 norm. We used the L2 norm loss to increase the semantic consistency between the attention maps *M*_*i,k*_ and SHAP priors SHAP_*k*_ corresponding to class *k* ∈ [1, *K*], thereby encouraging the model to focus its attention on disease-relevant regions that the pre-computed priors highlighted in the brain.

The final loss is then:

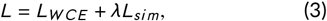

where *λ* is a hyper-parameter that can be optimized.

### 2.4 Neuropathological validation

To validate model predictions with gold standard biological evidence, we correlated deep feature contributions with region-specific neuropathological scores obtained from autopsy on persons who had their last MRI within a year of their demise. We quantified regional disease presence based on the degree of amyloid *β* deposits, neurofibrillary tangles (NFT), and neuritic plaques (NP) on histology. These three pathologies are hallmarks of AD that increase in density and/or spread through the brain as the disease progresses, and they are associated with tissue/cellular damage and death [37].

We obtained 23 participants from ADNI (*n* = 13) and FHS (*n* = 10) who had MRI scans taken within one year of death with available regional semi-quantitative histopathology scores. Presence and density of amyloid *β* deposits, neurofibrillary tangles, and neuritic plaques were assessed in the entorhinal, hippocampal, frontal, temporal, parietal, and occipital cortices. The regions were proposed based on the NIA-AA protocol for standardized neuropathological assessment of AD. Severity of the assessment was categorized into four score categories: 0 (None), 1 (Mild), 2 (Moderate), and 3 (Severe) [38]. We used the trained models to run inference on those cases and saved their corresponding class-wise attention maps for computation of region-level scores. Since postmortem histology grades assess for the presence of disease in the respective brain regions, we used the AD-specific attention map to compute region-level attention scores as model evidence for the prediction of AD. Using the MNI-152 template, we obtained a brain parcellation for each of the MRIs and aggregated voxel attention values per region, normalized by region volume. Once model scores were computed, we ran the Spearman’s rank correlation coefficient test with pathology grades of amyloid *β*, neurofibrillary tangles, and neuritic plaques in the various pre-identified brain regions.

## 3 EXPERIMENTAL SETUP

We considered the NACC dataset as the source domain for training, validation and internal testing, and used ADNI, AIBL, and FHS as the target domains for external testing. All experiments were run with *k* -fold cross validation over the source domain with *k* = 5, and the average metrics over the five runs with their standard deviation were reported. Since the source domain we have access to suffers from class imbalance, wherein MCI and AD cases are significantly less than NC cases, we used stratified *k* -fold cross validation to ensure the target classes follow the same ratio in each fold as in the full dataset. We used a split ratio of 3 : 1 : 1, where 60% of the data were used for model training, 20% were used for internal validation, and the rest for internal testing. We trained our models for 60 epochs with 200 steps, i.e., weight updates, per epoch. We also compared against two state-of-the-art methods in the single-source DG setting: RSC[15] and Mixup[16]. After hyper-parameter tuning, we chose a *λ* = 5 × 10^™5^ for our training strategy and an *α* = 0.2 for the Mixup[16] method. Due to large size of the input image, i.e., (182 × 218 × 182) per MRI, we could only fit a batch size of 2 into GPU memory (48 GB) and had to resort to gradient accumulation over 8 steps to simulate a final batch size of 16, since the small batch size rendered weighted random sampling ineffective for mitigating class imbalance. We also modified the state-of-the-art DG methods to use weighted cross-entropy across all experiments, as their implementation was not designed to deal with heavy class imbalance.

### 3.1 Performance metrics

Along with model accuracy, we reported the macro F1-score averaged over five folds as it better represents a balanced score between precision and recall through their harmonic mean. The macro F1-score in multi-class classification is the average of F1-scores over all classes (4). A higher macro F1 score represents lower false positives, i.e., recall, and false negatives, i.e., precision.

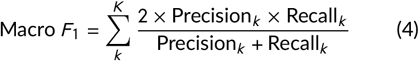

such that,

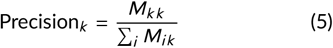

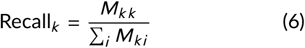

We also reported Matthew’s Correlation Coefficient (MCC), using Scikit-Learn’s [39] formula for multi-class classification (7). An advantage of having MCC as a single-value classification metric is that it is perfectly symmetric, unlike precision and recall that can be affected by swapping positive and negative classes. In addition, it quantifies how well the model is doing at predicting each class, regardless of class imbalance.

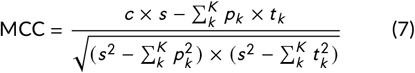

such that,

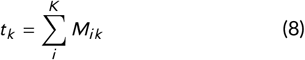

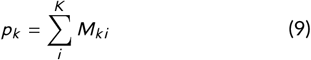

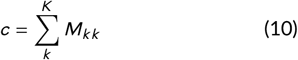

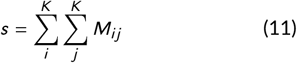

where *M* refers to the confusion matrix, *K* the total number of classes, *t*_*k*_ the number of times class *k* truly occured, *p*_*k*_ the number of times class *k* was predicted, *c* the total number of samples correctly predicted, and *s* the total number of samples.

### 3.2 Computing infrastructure

We used PyTorch (v1.13.1) and a NVIDIA A6000 graphics card with 48 GB memory on a GPU workstation to implement the model. The training speed was about 2.25s / iteration, and it took less than 24 hours to reach convergence with a batch size of 16 after gradient accumulation. The inference speed was < 0.2s per MRI.

### 3.3 Data and code availability

All the MRI scans and corresponding clinical and demographic data can be downloaded freely from ADNI, NACC and AIBL websites. FHS data is available upon request and subject to institutional approval. Python scripts and manuals are available on GitHub.^1^

## 4 RESULTS

We compared the results of our computational framework against state-of-the art DG methods for the single-source setting in Table 2. We used a vanilla UNet3D model trained without DG on the NACC cohort as the baseline on which we ran three different experiments: one trained from scratch and not using attention (row 1), another also trained from scratch but with our attention module (row 2), and the third trained starting from the pretrained `Models Genesis`[32, 33] check-point with our attention module (row 3). First, the two methods we compared against, RSC[15] and Mixup[16], did not show improvement over the baseline. In fact, performance was deteriorated going from row 3 to row 4 by 10.8% in terms of target mean accuracy, 0.07 (7%) in terms of target mean macro F1-score, and 0.08 (4%) in terms of target mean MCC. The same pattern of performance degradation was observed going from row 3 to row 5, with a 4.6% lower target mean accuracy, a 0.03 (3%) lower target mean macro F1-score, and 0.04 (2%) lower target mean MCC. These results imply that although these methods were proven to boost performance and robustness to distributional shifts on natural and synthetic imaging benchmarks, they do not translate to real-world clinical data, in our case, volumetric structural brain MRIs. On the other hand, training using our method improved performance, outperforming RSC[15], Mixup[16], and the baseline across the reported target mean metrics. We showed a 2.8% improvement over the baseline (row 3 vs. 7) in terms of target mean accuracy. In fact, our method was able to achieve a 73.4% accuracy on the target cohort AIBL, a 7.3% improvement over the baseline (row 7 vs. 3). This improvement is also reflected in the MCC value which increased by 0.07 (3%) from row 3 to row 7.

**T A B L E 2.**
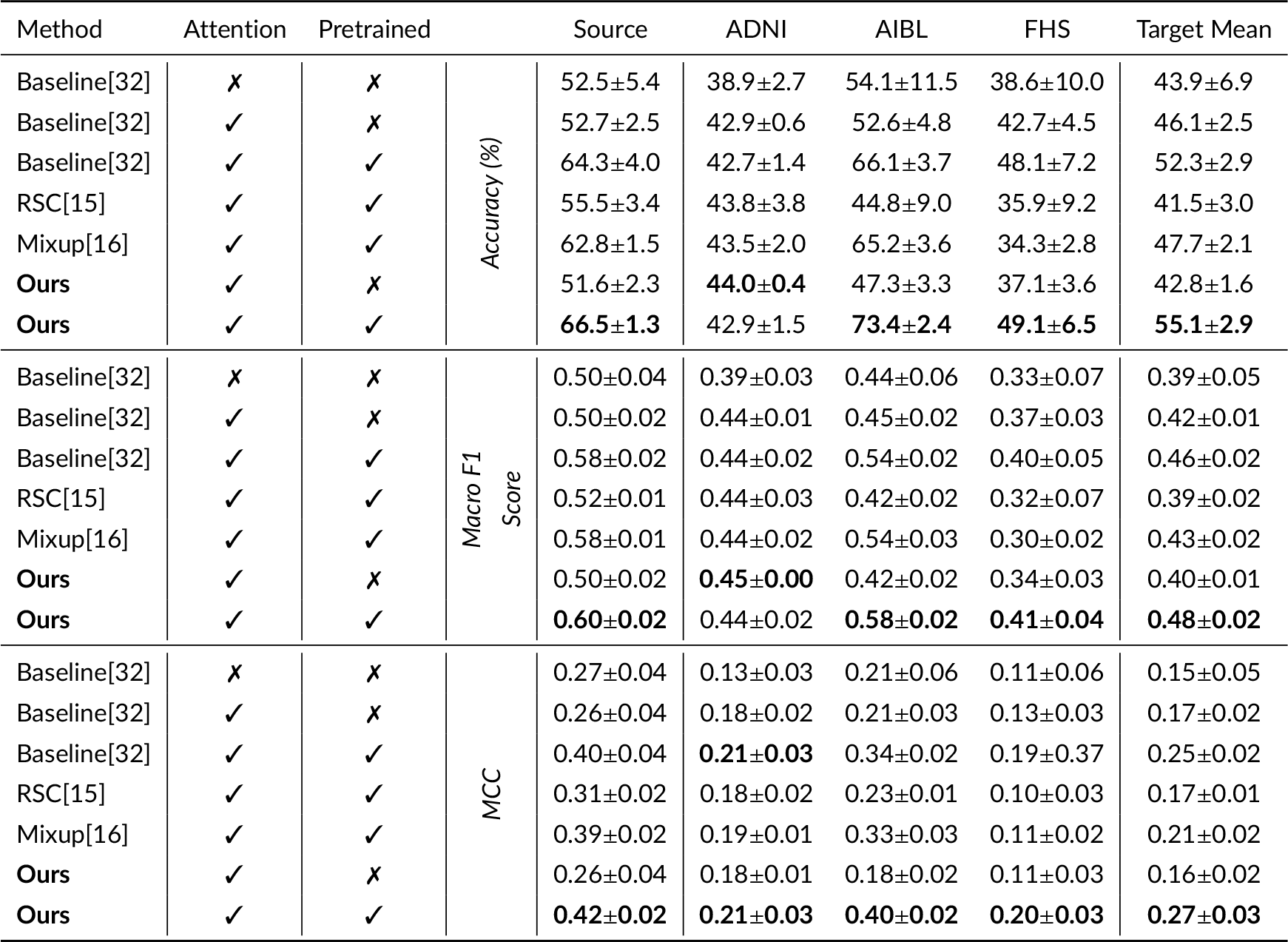
Model performance. We trained our model on the NACC cohort and used the ADNI, AIBL, and FHS cohorts as target domains. We reported accuracy on the test split of NACC, and on the entirety of the target datasets. Performance metrics including accuracy, macro F1-score and MCC are reported on each case. Note that model training was done via 5-fold cross validation on the NACC dataset, and testing was done on each of the models. Results are reported as mean ± standard deviation. The bold font is used to report the best model performance in each column.

The above quantitative results were reflected in the tSNE visualizations we presented in Fig. 3 of the baseline model trained without DG (row 3 in Table 2) and the model trained with our computational framework (row 7 in Table 2). While (a) shows the MRI embeddings learned by the baseline model clustered by cohort, (b) shows that our approach to aligning model attention with SHAP priors reduces cohort-specific clustering. In particular, the remarkable improvement in accuracy over the baseline on the AIBL cohort shows in the dispersion of MRI embeddings belonging to AIBL (green) across the tSNE plot in (b) as opposed to clear clusters highlighted by the red boxes in (a). These results indicate that even though the SHAP priors used in training were derived only from the source domain, they effectively reduced the distributional variance across source and target domains.

**F I G U R E 3.**
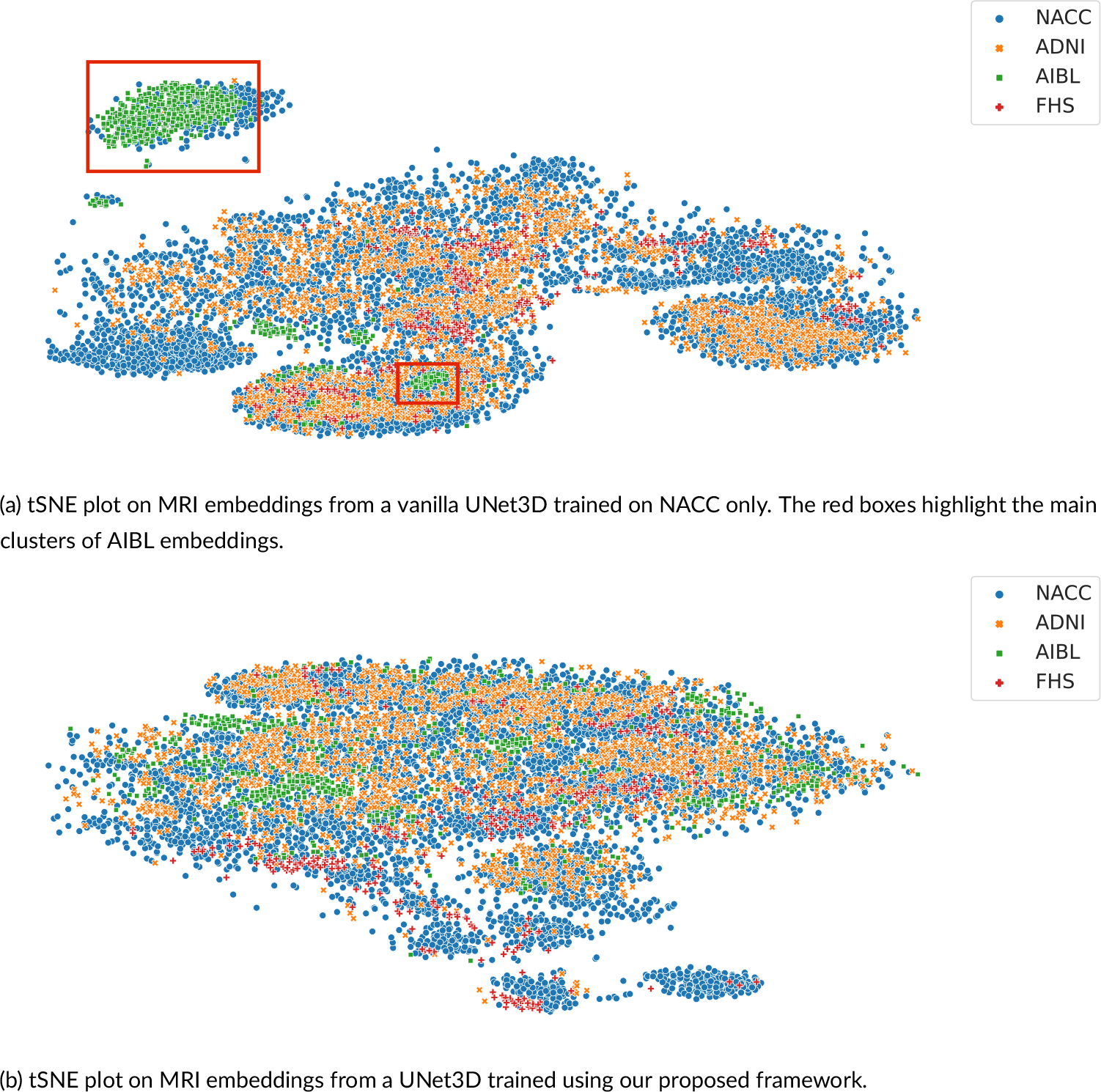
Visualization of MRIs in the latent space. (a) We generated MRI embeddings at the attention module level from a vanilla UNet3D model trained on the NACC cohort without domain generalization, and visualized them in a 2D space using t-SNE. (b) We generated MRI embeddings from the same UNet3D architecture trained using our domain generalization framework, and visualized them in a 2D space using t-SNE. For both plots, data from the four cohorts (NACC (*n* = 4, 647), ADNI (*n* = 1, 821), AIBL (*n* = 661) and FHS (*n* = 304)) were used.

We further validated our method with gold standard evidence of disease pathology and compared it against the other methods, reporting the results in the form of a correlation heat map in Fig. 4. We showed that not only did our method correlate more strongly with postmortem histology scores than other methods, but also, our results were more consistent across the three stains. Correlation of our method with pathology in the amygdala, hippocampus, parahippocampal and ambient gyri was positive for amyloid *β*, neurofibrillary tangles, and neuritic plaques. We then projected the computed correlation values onto their corresponding brain regions and displayed the projections (Fig. 5). Subfigure (a) shows an improved correlation for our method with pathology grades of amyloid *β* in the hippocampal region and the middle frontal gyrus of the frontal lobe. Correlation in these brain regions is also consistent with pathology grades of neurofibrillary tangles and neuritic plaques (subfigures (b) and (c)). As for the other evaluated methods, shown in the first three columns of each subfigure, the correlations were lower with pathology grades in the hippocampus of amyloid *β*, neurofibrillary tangles, and neuritic plaques, except for the baseline method in subfigure (c) that had a positive - although lower than ours - correlation. In addition, our method showed the highest correlation in the parahippocampal and ambient gyri with pathology grades of neuritic plaques in subfigure (c). Our method demonstrated high correlations with specific brain regions, notably the hippocampal and parahippocampal areas, which were visually represented in the pre-computed AD-specific SHAP priors (Fig. 2). These regions contributed positively to model predictions of AD, indicating the effectiveness of our technique in aligning model attention with established knowledge regarding disease indicators. Such observations indicating improved model correlation with regions that are well-known to be implicated with disease grounded our model predictions with biological evidence.

**F I G U R E 4.**
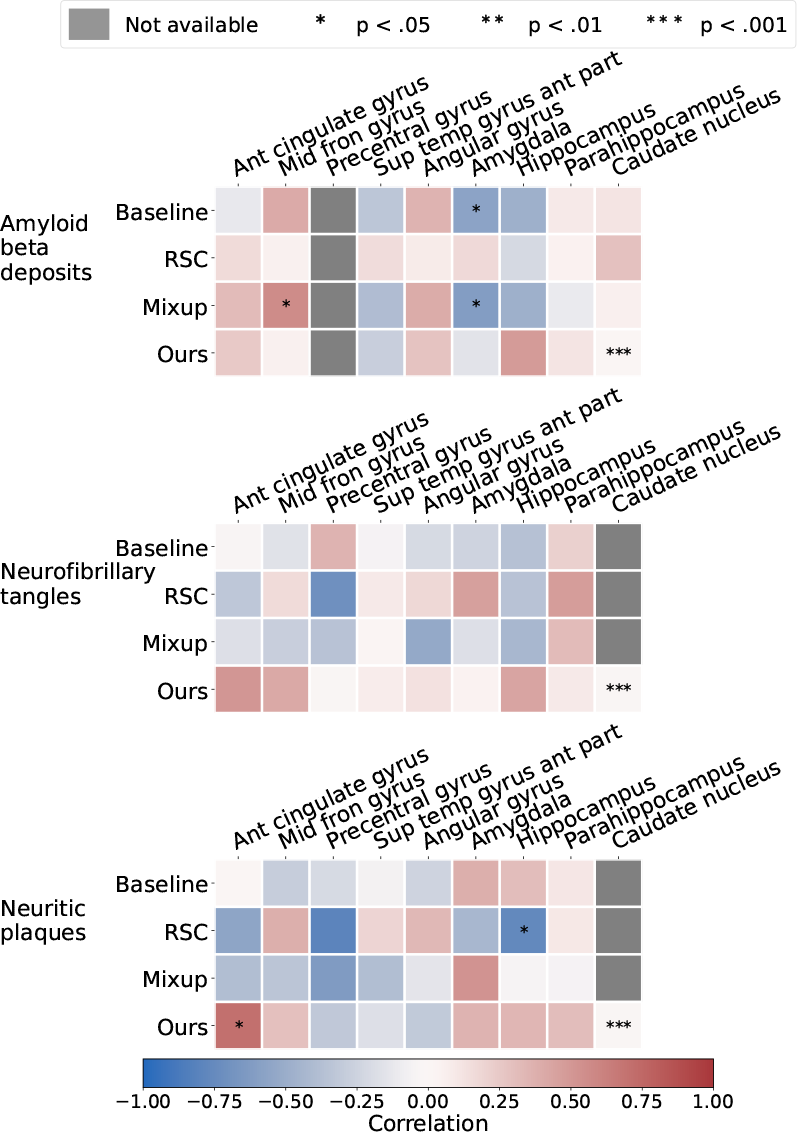
Correlation of model-generated attention scores with post-mortem histology. Pathology grades of amyloid *β*, neurofibrillary tangles and neuritic plaques in various brain regions on deceased ADNI and FHS participants were obtained (*n* = 23). We compared model-identified importance in these brain regions with the degree of pathology severity, and compared them against predictions obtained using other well-known domain generalization methods.

**F I G U R E 5.**
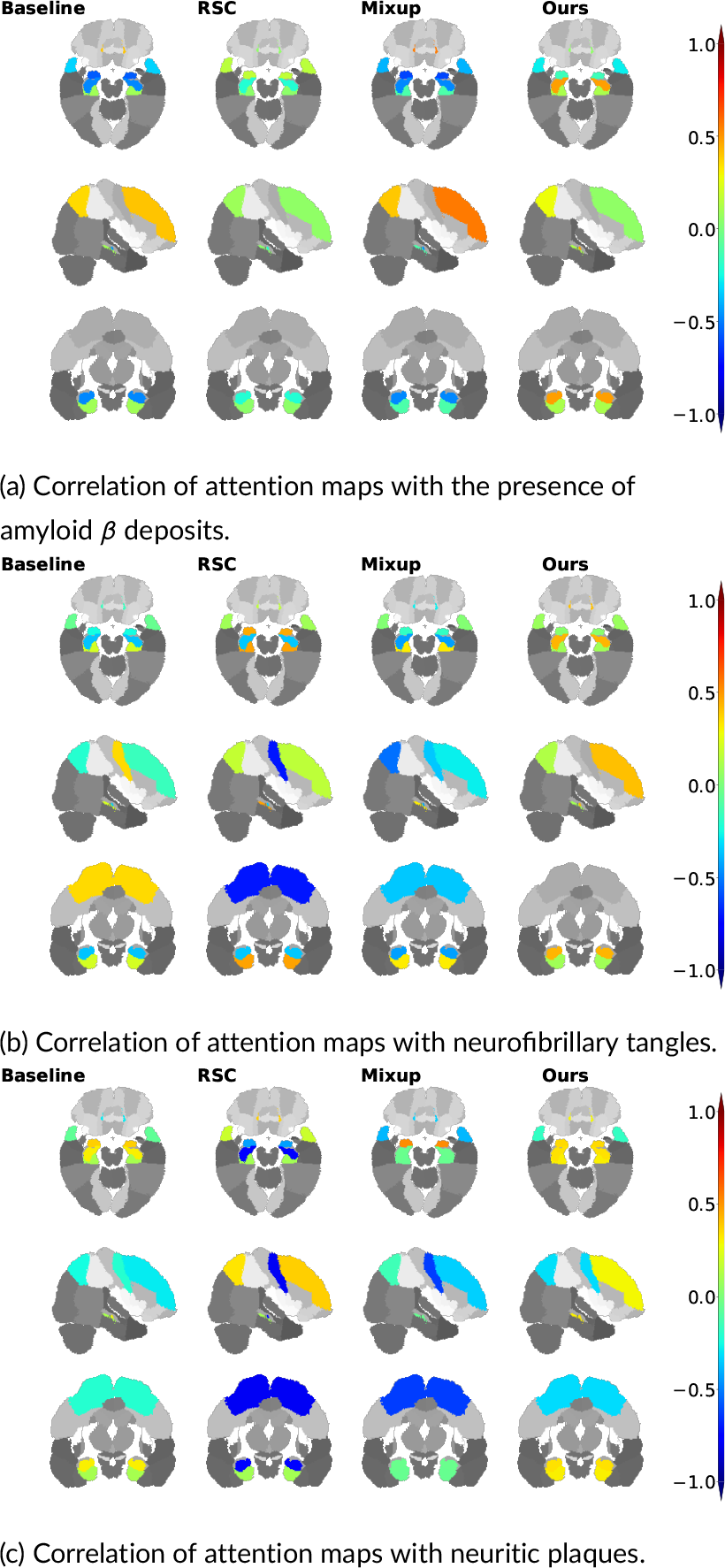
Visualization of correlations between model attention scores and post-mortem histology. We obtained region-specific pathology grades of amyloid *β*, neurofibrillary tangles and neuritic plaques on deceased ADNI and FHS participants (*n* = 23). We computed Spearman’s rank correlation coefficient between model-derived attention scores and the region-specific pathology grades and projected them on the corresponding brain regions.

## 5 DISCUSSION

This work presents a computational framework for DG that adds disease-driven interpretability to deep learning models for Alzheimer’s disease prediction on volumetric MRI scans. While most of the existing methods focus on achieving high model performance on unseen data, they do not directly account for the underlying disease biology during model development. We achieved this goal by refining the model’s attention to focus on brain regions that are most associated with disease based on pre-computed feature contributions. In such fashion, our method distinguishes itself by incorporating disease-driven interpretability into the training process. The added interpretability can provide a better understanding of the underlying disease mechanisms and aid in the clinical decision-making process. We compared the performance of our method with previously published DG frameworks, and showed that our approach shows competitive performance while incorporating disease relevance into the model training process. We confirmed the degree to which our attention-based supervision strategy ultimately reflected disease biology by comparing model attention in predefined brain regions with postmortem neuropathology scores. Overall, our approach to creating a generalizable framework complements other published work in the literature.

We observed that our model achieved consistent, favorable performance on the test cohorts relative to other DG frameworks. While extensive testing is required to confirm any modeling framework’s superiority in accurate prediction of disease, it is worth noting that model performance based on accuracy alone without downstream evidence of correlation with a reference standard may not be appealing in the context of medical machine learning. As such, classifying persons with NC from those who have MCI or AD solely on MRIs is a clinically challenging task, and often not part of routine clinical neurology work-up. Neurologists use a spectrum of patient data along with MRIs to assess a person’s cognitive status. Nevertheless, our proposed framework has utility in the objective interpretation of brain MRIs, and broadly in the quantification of findings indicative of disease. Besides minimizing subjectivity, it also potentially fills gaps in healthcare settings where there is a lack of neuroradiology expertise.

Our study has a few limitations. Due to memory limitations, we resorted to offline computation of the saliency maps based on correct predictions by the trained baseline model. We also acknowledge that SHAP prior computation is solely dependent on the baseline model used, i.e., the quality of prior knowledge and correctness of feature contributions extracted from the model are directly correlated with model performance. Also, it is possible that the offline computation and aggregation of class-specific SHAP maps may have reduced instance-to-instance variability and minimized radiologic artifacts, thereby facilitating model attention on disease pathology. In addition, it is possible that the model was able to capture the fine-grained nature of disease markers due to our choice of the voxel-wise L2 distance metric. We utilized this loss function to increase the semantic similarity between model attention and prior maps at the voxel level.

In conclusion, our work contributes to the growing field of interpretable deep learning in medical imaging, paving the way for more accurate and personalized diagnoses of cognitive disorders. By highlighting the specific brain regions that contribute most significantly to disease, our approach can provide valuable insight into disease mechanisms and aid in developing targeted interventions. Furthermore, the disease-driven interpretability of our framework can help build trust and understanding between clinicians and patients, which is crucial for effective healthcare delivery.

## Data Availability

All the MRI scans and corresponding clinical and demographic data can be downloaded freely from ADNI, NACC and AIBL websites. FHS data is available upon request and subject to institutional approval.

https://naccdata.org

https://adni.loni.usc.edu

https://adni.loni.usc.edu/category/aibl-study-data/

## 6 ACKNOWLEDGEMENTS

This project was supported by grants from the Karen Toffler Charitable Trust, the American Heart Association (20SFRN35460031), the National Institutes of Health (RF1-AG062109, R01-HL159620, R21-CA253498, and R43-DK134273), and a pilot award from the National Institute on Aging’s Artificial Intelligence and Technology Collaboratories (AITC) for Aging Research program.

## 7 AUTHOR CONTRIBUTIONS

DL, SAB, BAP, and VBK: study conception and design. DL and SS: data collection and processing. DL: implementation. DL and SS: analysis. DL, SS, SAB, BAP, RA and VBK: data interpretation and manuscript write-up. VBK: study direction. All authors reviewed the results and approved the final version of the manuscript.

## Footnotes

1 https://github.com/vkola-lab/d3g.git

